# Duration and reliability of the silent period in individuals with spinal cord injury

**DOI:** 10.1101/2020.06.07.20124701

**Authors:** Hannah Sfreddo, Jaclyn R. Wecht, Ola Alsalman, Yu-Kuang Wu, Noam Y. Harel

## Abstract

**Designs:** Observational.

**Objectives:** We aim to better understand the silent period (SP), an inhibitory counterpart to the well-known motor evoked potential (MEP) elicited by transcranial magnetic stimulation (TMS), in individuals with spinal cord injury (SCI).

**Setting:** Veterans Affairs Hospital in New York.

**Methods:** Electromyographic responses were measured in the target abductor pollicis brevis at rest (TMS at 120% of resting motor threshold (RMT)) and during maximal effort (TMS at 110% of RMT). Participants with chronic cervical SCI (n=9) and able-bodied volunteers (n=12) underwent between 3-7 sessions of stimulation on separate days. The primary outcomes were the magnitude and reliability of SP duration, resting and active MEP amplitudes, and RMT.

**Results:** SCI participants showed significantly lower MEP amplitudes compared to AB participants. SCI SP duration was not significantly different from AB SP duration. SP duration demonstrated reduced intra-participant variability within and across sessions compared with MEP amplitudes. SCI participants also demonstrated a higher prevalence of SP ‘interruptions’ compared to AB participants.

**Conclusions:** SP reflects a balance between corticospinal excitatory and inhibitory processes. SP duration is more reliable within and across multiple sessions than MEP amplitude.

## 1 Introduction

The motor evoked potential (MEP) amplitude in response to transcranial magnetic stimulation (TMS) is one of the most commonly used outcome measures for tracking neurophysiology of the corticospinal system. Spinal cord injury (SCI) reduces MEP amplitude in muscles below the injury, reflecting reduced excitatory transmission across the spinal lesion. In contrast, corticospinal inhibitory processes are comparatively understudied after SCI. The silent period (SP) occurs upon TMS administration to the motor cortex during volitional contraction of a target muscle on either the ipsilateral or contralateral side of stimulation. In contralateral SP elicitation, the resulting MEP is followed by a period of electromyographic (EMG) suppression (silent period) usually lasting 100-300 milliseconds^1–3^ (Figure 1). Whereas the ipsilateral SP depends on corpus callosum-mediated interhemispheric inhibition, the contralateral SP reflects cortical and corticospinal processes^3,4^.

**Figure 1.**
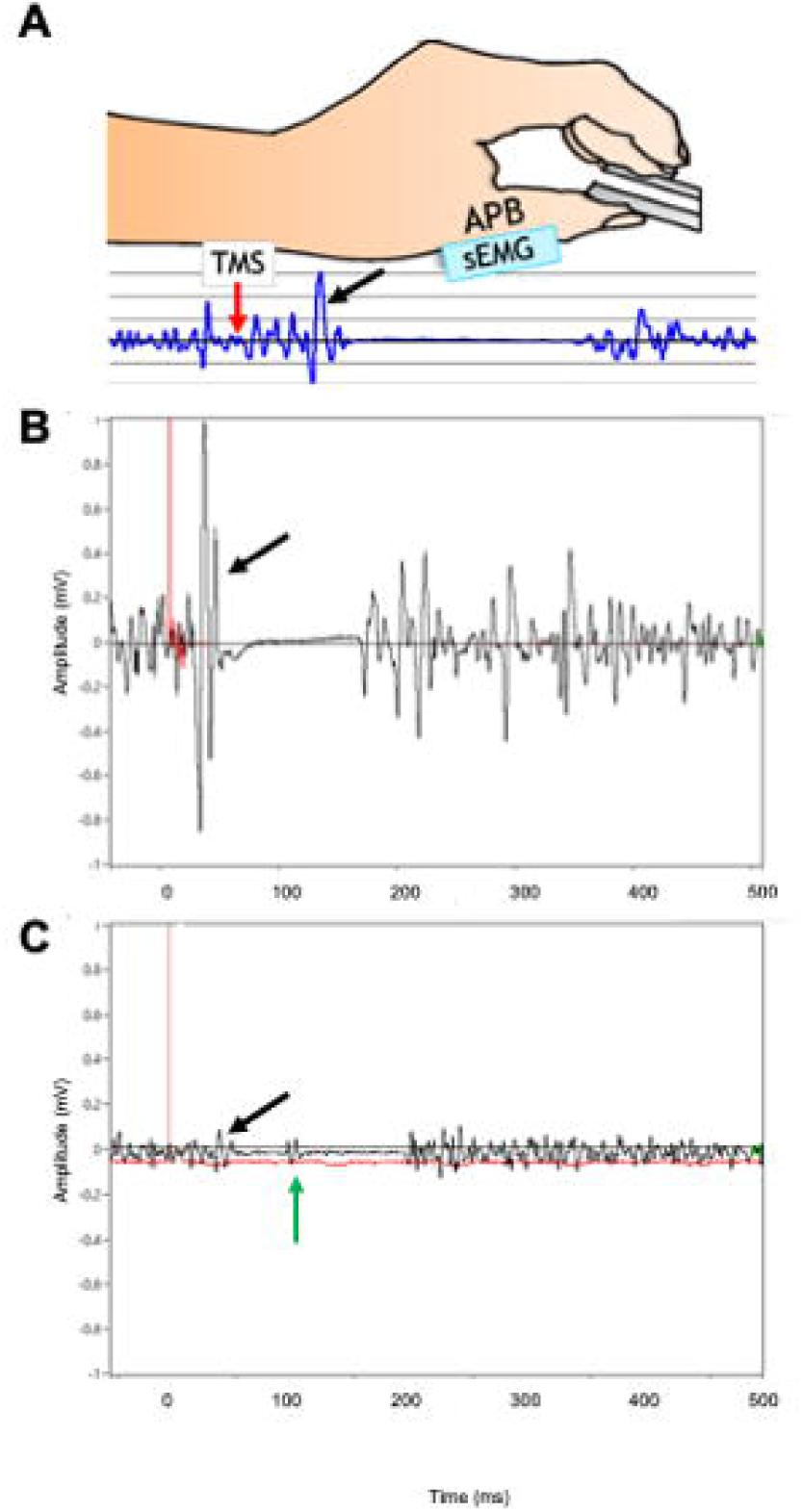
Schematic and examples of silent period. **A)** TMS is delivered (red arrow) to the hand motor cortex while the participant performs a volitional isometric pinch with the contralateral hand. The resulting MEP (black arrow) is followed by an EMG silent period before the resumption of baseline volitional EMG activity. **B) Representative silent period in able-bodied volunteer. C) Representative silent period in SCI participant**. Note lower EMG amplitudes and presence of ‘interruption’ (green arrow). APB, abductor pollicis brevis.

Spinal inhibitory mechanisms contribute to the first ∼50 ms of the SP through hyperpolarization and segmental recurrent inhibition of motor neurons^1,3^. Interneuronal gamma-aminobutyric acid (GABA)_B_ plays a major role in mediating intracortical inhibition of pyramidal motor cortex neurons during the remainder of the SP^2,5^. This is most clearly demonstrated by the elongation of SP duration when baclofen, a specific GABA_B_ receptor agonist, is delivered intrathecally^6^. However, studies of oral and intravenous baclofen have failed to demonstrate a significant effect on SP duration^5,7,8^.

Lesions of the nervous system may lengthen or shorten SP duration. Cerebral pathologies that lengthen SP include stroke^9^, epilepsy^10^, and depression^11^. Conversely, pathologies that shorten SP include bipolar disorder^12^ and chronic neuropathic pain^13^.

Few prior studies have investigated SP in the SCI population, with mixed results^7,14–17^. It is not yet fully elucidated whether SCI lengthens SP (perhaps due to decreased afferent feedback from the periphery) or shortens SP (perhaps due to increased cortical or spinal segmental excitability). Furthermore, the reliability of SP duration as an outcome metric in SCI relative to the more commonly used MEP amplitude is hardly known^18^. To further characterize this potentially important neurophysiological measure in the SCI population, we compared magnitude and variability of hand muscle SP durations, MEP thresholds, and MEP amplitudes in individuals with chronic cervical SCI and able-bodied volunteers across multiple testing sessions.

## 2 Methods

### 2.1 Design

This experiment was an exploratory post-hoc analysis of a larger study in which we tested a novel configuration for non-invasive cervical transcutaneous spinal stimulation in individuals with and without cervical SCI (clinicaltrials.gov NCT02469675)^19^. SCI participants were eligible if they had traumatic or non-traumatic SCI between segments C2-C8 with any evidence for partially retained movement of finger extension, finger flexion, or finger abduction of either hand. Potential participants were excluded if they had risk factors for seizures or if they had frequent episodes of autonomic dysreflexia. Participants provided informed consent before initiating testing. All procedures were approved by the Institutional Review Board of the James J. Peters VA Medical Center, Bronx, NY. All applicable institutional and governmental regulations concerning the ethical participation of human volunteers were followed during this research.

### 2.3 General Procedure

Neurological examination of motor and sensory function was performed according to the International Standards for the Neurological Classification of Spinal Cord Injury (ISNCSCI). Sessions were performed on separate days at a consistent time of day per participant. Stimulation was delivered with participants in a seated upright position in an adjustable TMS treatment chair (Magventure), or for one participant, in her own wheelchair. For participants without neurological injury, TMS was targeted toward the dominant hand. For those with SCI, TMS was targeted toward the hand with lower motor thresholds and more consistent electrophysiological responses to central and peripheral stimulation. Arms and hands were pronated and relaxed on a cushion placed in the participant’s lap. Participants were not asked to withhold their routine daily medications.

### 2.4 Transcranial Magnetic Stimulation (TMS)

A MagPro R30 or X100 system (Magventure, Farum Denmark) with 80mm winged coil (D-B80) was used. The magnet was oriented at a 45-degree angle from the sagittal plane, centered over the hand motor cortex hotspot for maximal APB response. The first six participants wore reusable cloth headcaps upon which the hotspot was labeled with a marker in relation to the vertex. Our laboratory obtained an optical-based neural navigation system (Brainsight, Rogue Research, Montreal, Canada) that was used to track hotspots for the final 15 participants. There was no significant difference in session-to-session variability of any TMS measure with or without neural navigation. RMT was determined as the percent of maximal stimulator output required to elicit an MEP in the APB muscle of at least 50 μV in 5 out of 10 repetitions.

### 2.5 Electromyographic Data

EMG of the target APB was recorded using surface sensors with 300x preamplification, 15-2,000 Hz bandwidth, and internal grounding (Motion Lab Systems Z03-002, Baton Rouge, Louisiana, USA). EMG was collected at a sample rate of 5,000 Hz via digital acquisition board and customized LabVIEW software (National Instruments USB-6363, Austin, Texas, USA). All EMG data were acquired and quantified using custom LabVIEW scripts.

### 2.6 Eliciting and Measuring the Silent Period (SP)

While participants pinched a dynamometer (Tracker Freedom, J-Tech, Salt Lake City, Utah, USA) between their thumb and third finger using maximal effort, a single biphasic TMS pulse was delivered over the hand motor cortex hotspot at 110% of each participant’s RMT. The resulting MEP amplitude and SP duration in the contralateral APB muscle were measured. Five to six replicates were performed per session, with care taken to avoid fatigue between replicates. Participants completed 7 sessions on different days involving SP, apart from three individuals who withdrew from the study before completion (one AB participant completed 6 sessions, one SCI participant completed 3 sessions, and one SCI participant completed 2 sessions).

To quantify SP_DUR_, SP onset was visually defined as the end of the TMS-induced MEP, and SP offset was visually defined as the earliest resumption of pre-TMS EMG activity^3^ (Figure 1).

We defined an “SP interruption” as a spike in EMG activity surrounded by SP silence, where the duration of interruption was less than 20 ms, and the absolute amplitude from one peak to the adjacent trough was at least 25% of the largest amplitude found during the 100 ms preceding the TMS impulse.

### 2.7 Analysis

#### Outcomes

The primary dependent variables were MEP amplitude (mV) at rest (MEP_REST_) and during SP elicitation (MEP_ACTIVE_), SP duration (ms) (SP_DUR_), and RMT (% maximum stimulator output). SP interruptions were an exploratory outcome. For each participant, within-session means were computed from the 5-6 replicates of each test, then averaged across sessions. Before group comparisons, each outcome was first tested for normality using the Shapiro-Wilks test. Normally distributed values (MEP_ACTIVE_, SP_DUR_, and RMT) were compared between AB and SCI groups using independent-sample t-tests. Non-normally distributed values (MEP_REST_) were compared using an independent-samples Mann-Whitney U test.

#### Variability

Within-session coefficients of variation (CV) were computed for each participant from the 5-6 replicates of each test, then averaged across sessions^20^. Note that RMT was only determined once per session, so it was not possible to determine within-session RMT variability. Between-session CVs were computed from the session means of each test. CVs were then averaged across participants within each group. As CV values were not normally distributed, the non-parametric related-samples Friedman’s two-way analysis of variance by ranks test was applied to compare CVs across outcomes within each group. Significant values on Friedman’s test were analyzed post-hoc between pairs of outcomes using related-samples Wilcoxon signed rank tests.

#### Interruptions

A chi-squared test was used to compare the frequency of participants with interrupted SPs between groups. As interruption values were not normally distributed, the non-parametric Mann-Whitney U test was used to compare number of interruptions per participant between groups.

Due to testing multiple comparisons, a p value of <0.01 was used to determine significance.

Excel (Microsoft, Redmond, Washington, USA) and SPSS Version 25 (IBM, Armonk, New York, USA) were used for all analyses.

Individual-level data is included as Supplementary Data 1 (by participant, averaged across sessions) and Supplementary Data 2 (by participant per session).

## 3 Results

### 3.1 Participants

21 individuals (12 AB and 9 SCI; 17 males, 4 females) participated in this study (Table 1). Participants ranged in age from 22 to 64 years old. Of the 9 SCI participants, eight had traumatic SCI, one had idiopathic transverse myelitis.

**Table 1.**
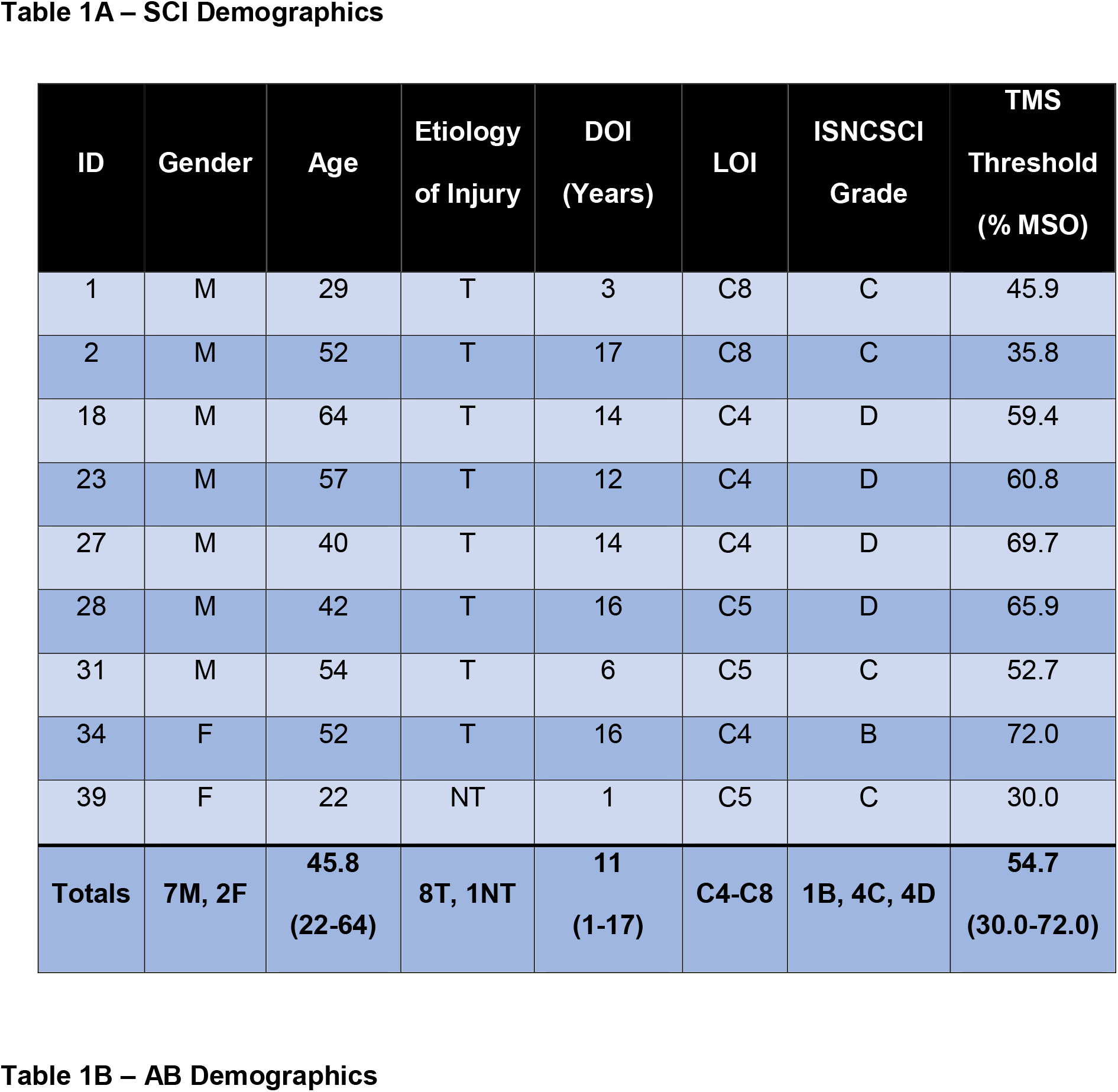
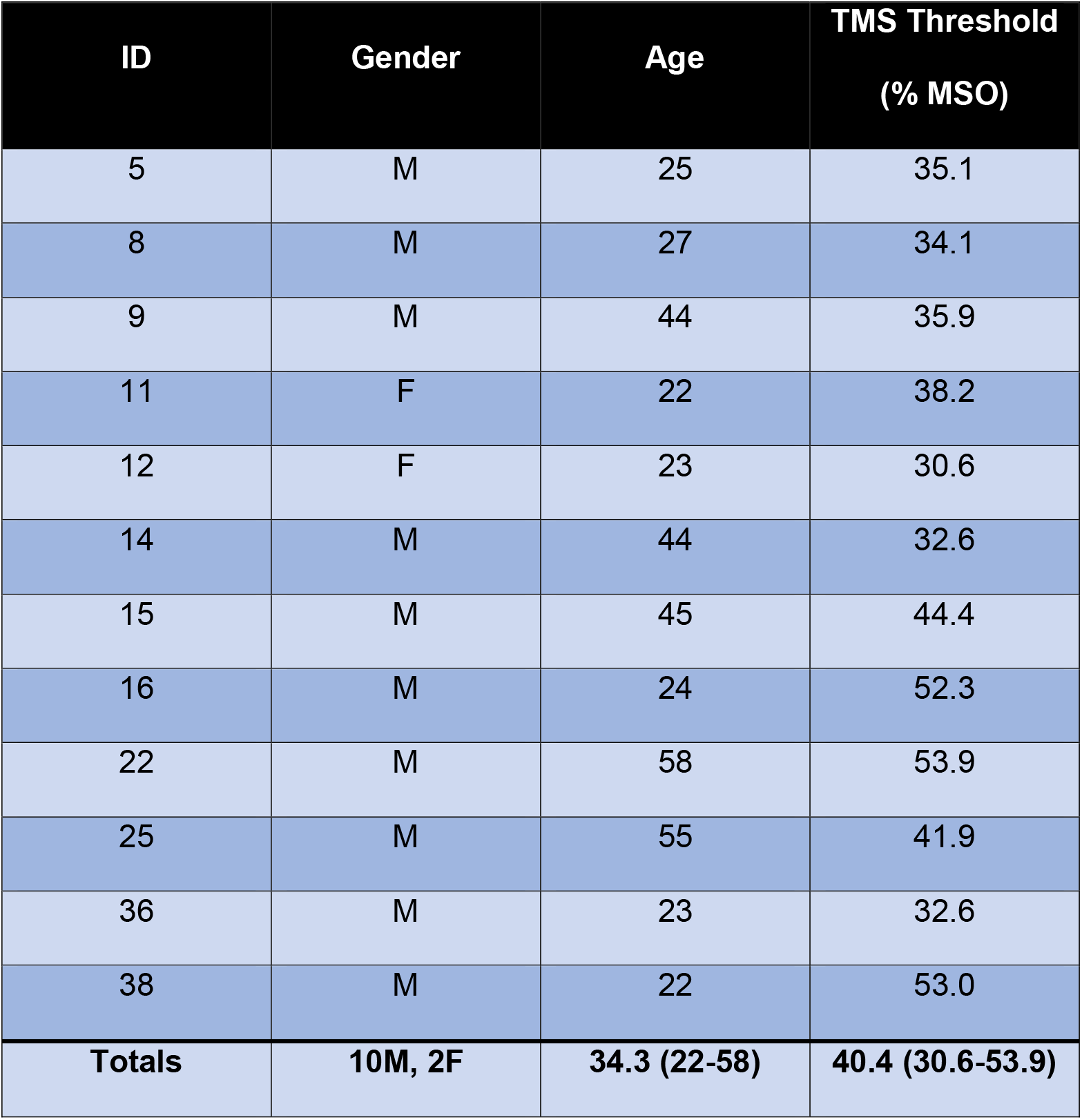
Participant demographics. **A) SCI**. Etiology of Injury – traumatic (T) or not traumatic (NT); DOI – duration of injury; LOI – neurological level of injury; ISNCSCI – International Standards for the Neurological Classification of SCI; %MSO – percent of maximal stimulator output (average across sessions). **B) AB**.

### 3.2 SP duration

SCI participants had mean (SEM) SP duration of 111.40 (18.39) ms; range 46.90 – 216.17 ms, whereas AB participants had SP duration of 97.99 (9.88) ms; range 35.30 – 152.86 ms (non-significant, independent-sample t-test) (Table 2).

**Table 2.**
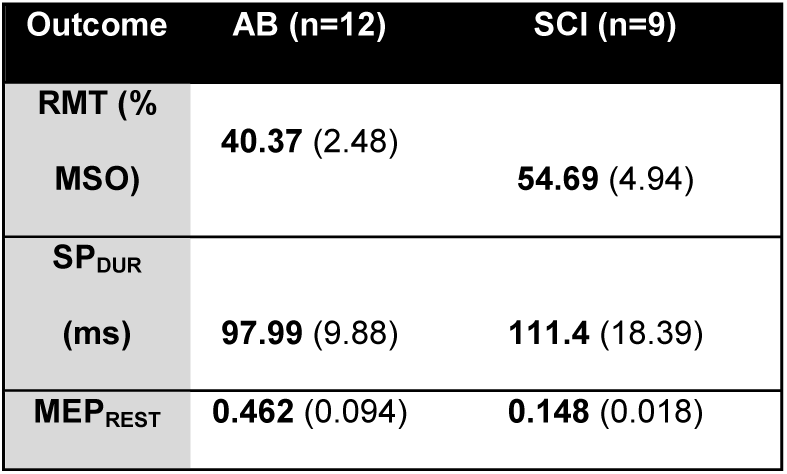
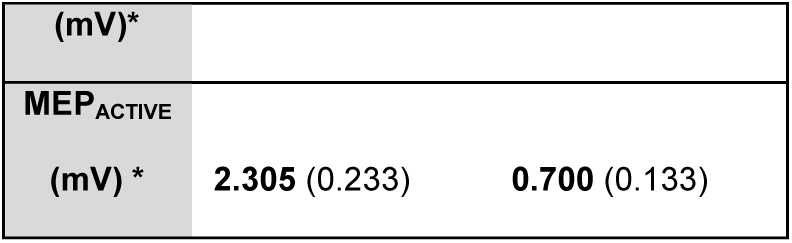
Between-group comparisons. Means across sessions (S.E.M.). RMT, resting motor threshold. % MSO, percent maximum stimulator output. SP_DUR_, SP duration. ms, milliseconds. MEP_REST_, amplitude of resting motor evoked potential. mV, millivolts. MEP_ACTIVE_, amplitude of active motor evoked potential. *, statistically significant between-group difference.

### 3.3 Resting motor threshold

SCI participants showed a tendency toward higher mean (SEM) RMTs (54.69 (4.94)% maximum stimulator output; range 30.0 – 72.0) than AB participants (40.37 (2.48)% maximum stimulator output, range 30.6 – 53.9) (p=0.012, independent-sample t-test) (Table 2).

### 3.4 MEP amplitudes

SCI participants showed significantly lower mean (SEM) MEP_REST_ amplitudes (0.148 (0.018) mV) than AB participants (0.462 (0.094) mV) (p<0.0005 independent-samples Mann-Whitney U test). SCI participants showed significantly lower MEP_ACTIVE_ amplitudes (0.700 (0.133) mV) than AB participants (2.305 (0.233) mV) (p<0.0005 independent-sample t-test) (Table 2).

### 3.5 Intraindividual variability within sessions (Table 3, Figure 2A)

In AB volunteers, the within-session CV (SEM) was 13.05 (1.40) for SP_DUR_, 59.79 (4.49) for MEP_REST_, and 21.83 (1.78) for MEP_ACTIVE_ (p<0.0005 on related-samples Friedman’s two-way analysis of variance by ranks test). Post hoc pairwise comparisons showed that SP_DUR_ had significantly lower within-session CV than both MEP_REST_ (p=0.002 on related-samples Wilcoxon signed rank test) and MEP_ACTIVE_ (p=0.005 on related-samples Wilcoxon signed rank test). In SCI participants, the CV was 16.08 (2.56) for SP_DUR_, 52.93 (6.18) for MEP_REST_, and 28.38 (2.64) for MEP_ACTIVE_ (p<0.0005 on related-samples Friedman’s two-way analysis of variance by ranks test). Post hoc pairwise comparisons showed that SP_DUR_ had significantly lower within-session CV than MEP_REST_ (p=0.008 on related-samples Wilcoxon signed rank test) but not MEP_ACTIVE_ (p=0.028 on related-samples Wilcoxon signed rank test). Thus, SP_DUR_ has lower within-session variability than MEP amplitudes in both able-bodied and SCI individuals. Variability did not significantly differ between AB and SCI groups.

**Table 3.** Outcome variability. Coefficients of variation (S.E.M.) for AB and SCI groups. **\***, statistically significant difference between SP_DUR_ and other outcomes within each group. **‡**, statistically significant difference between RMT and other outcomes within each group.

| Outcome | Group | Within-session | Between-session |
| --- | --- | --- | --- |
| RMT | AB |  | 6.96 (0.69) |
|  | SCI |  | 9.19 (1.21) |
| SP<sub>DUR</sub> | AB | 13.05 (1.40) | 19.61 (2.67) ‡ |
|  | SCI | 16.08 (2.56) | 28.50 (5.30) ‡ |
| MEP<sub>REST</sub> | AB | 59.79 (4.49) * | 52.04 (6.32) *‡ |
|  | SCI | 52.93 (6.18) * | 45.08 (6.90) |
| MEP<sub>ACTIVE</sub> | AB | 21.83 (1.78) * | 33.58 (4.12) ‡ |
|  | SCI | 28.38 (2.64) | 35.06 (3.63) ‡ |

**Figure 2.**
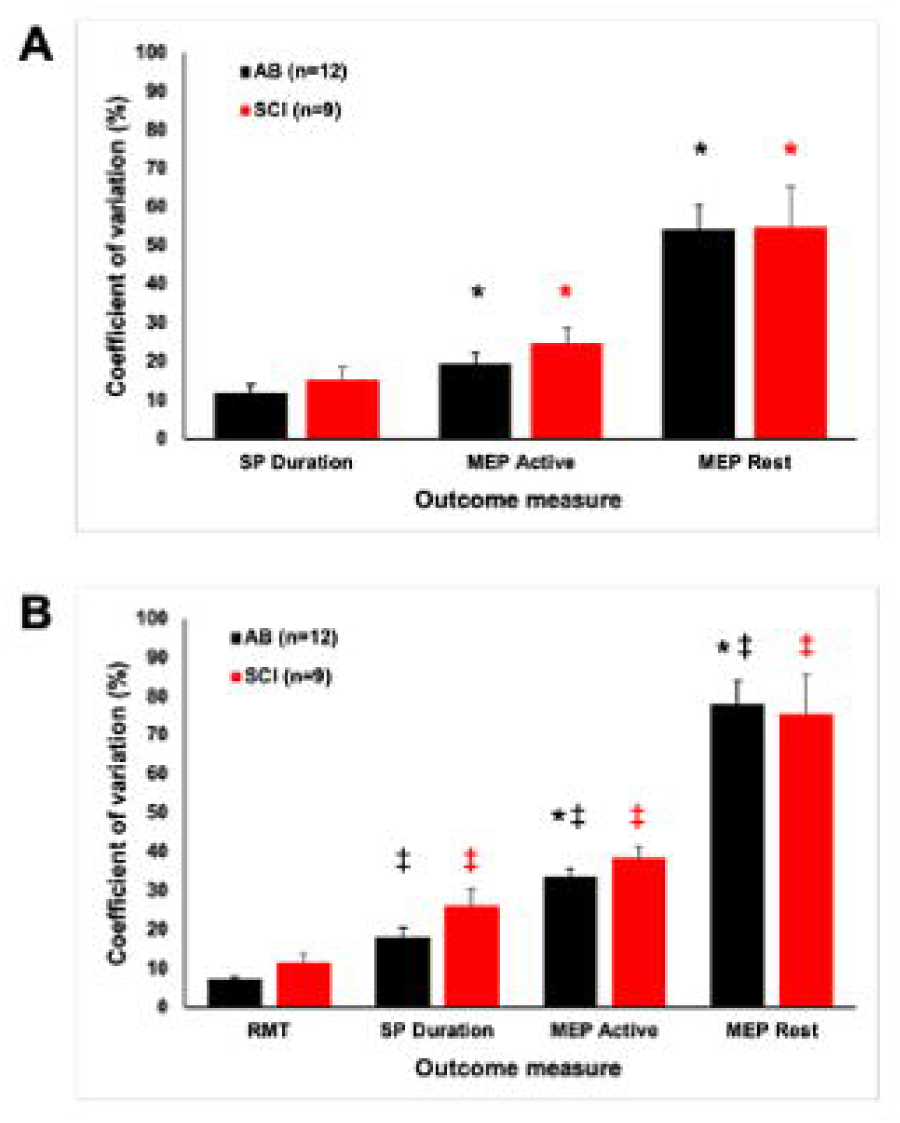
Outcome variability. **A)** Within-session coefficient of variation for each group. **B)** Between-session coefficient of variation for each group. Error bars represent S.E.M. *, significantly different from SP duration; ‡, significantly different from RMT.

### 3.6 Intraindividual variability across sessions (Table 3, Figure 2B)

In AB volunteers, the between-session CV was 19.61 (2.67) for SP_DUR_, 52.04 (6.32) for MEP_REST_, 33.58 (4.12) for MEP_ACTIVE_, and 6.96 (0.69) for RMT (p<0.0005 on related-samples Friedman’s two-way analysis of variance by ranks test). Post hoc pairwise comparisons showed that SP_DUR_ had significantly lower between-session CV than MEP_REST_ (p=0.002 on related-samples Wilcoxon signed rank test) but not MEP_ACTIVE_ (p=0.019 on related-samples Wilcoxon signed rank test), and significantly higher between-session CV than RMT (p=0.004 on related-samples Wilcoxon signed rank test). In SCI participants, the CV was 28.50 (5.30) for SP_DUR_, 45.08 (6.90) for MEP_REST_, 35.06 (3.63) for MEP_ACTIVE_, and 9.19 (1.21) for RMT (p<0.0005 on related-samples Friedman’s two-way analysis of variance by ranks test). Post hoc pairwise comparisons showed that SP_DUR_ had a non-significant trend toward lower between-session CV than MEP_REST_ (p=0.086 on related-samples Wilcoxon signed rank test), and no significant difference from MEP_ACTIVE_ (p=0.314 on related-samples Wilcoxon signed rank test). SP_DUR_ had a trend toward higher between-session CV than RMT (p=0.015 on related-samples Wilcoxon signed rank test) in SCI participants. There was no difference in between-session CV between AB and SCI participants. Thus, SP_DUR_ had significantly lower between-session variability than MEP amplitudes in able-bodied but not SCI individuals. However, RMT values were clearly more reliable across sessions than SP duration or MEP amplitudes.

### 3.7 SP Interruptions

3 out of 12 AB participants and 8 out of 9 SCI participants showed interrupted SPs (p =0.004; Pearson chi-square test). Per session, AB volunteers showed 0.089 (0.71) interruptions, while SCI volunteers showed 1.781 (0.787) interruptions (p=0.001 on independent-samples Mann-Whitney U test) (Figure 3). From the perspective of individual SP trials, there were five interrupted CSPs out of 467 total CSP events (1.07%) in AB participants, and 52 interrupted CSPs out of 242 total CSP events (21.5%) in SCI participants.

**Figure 3.**
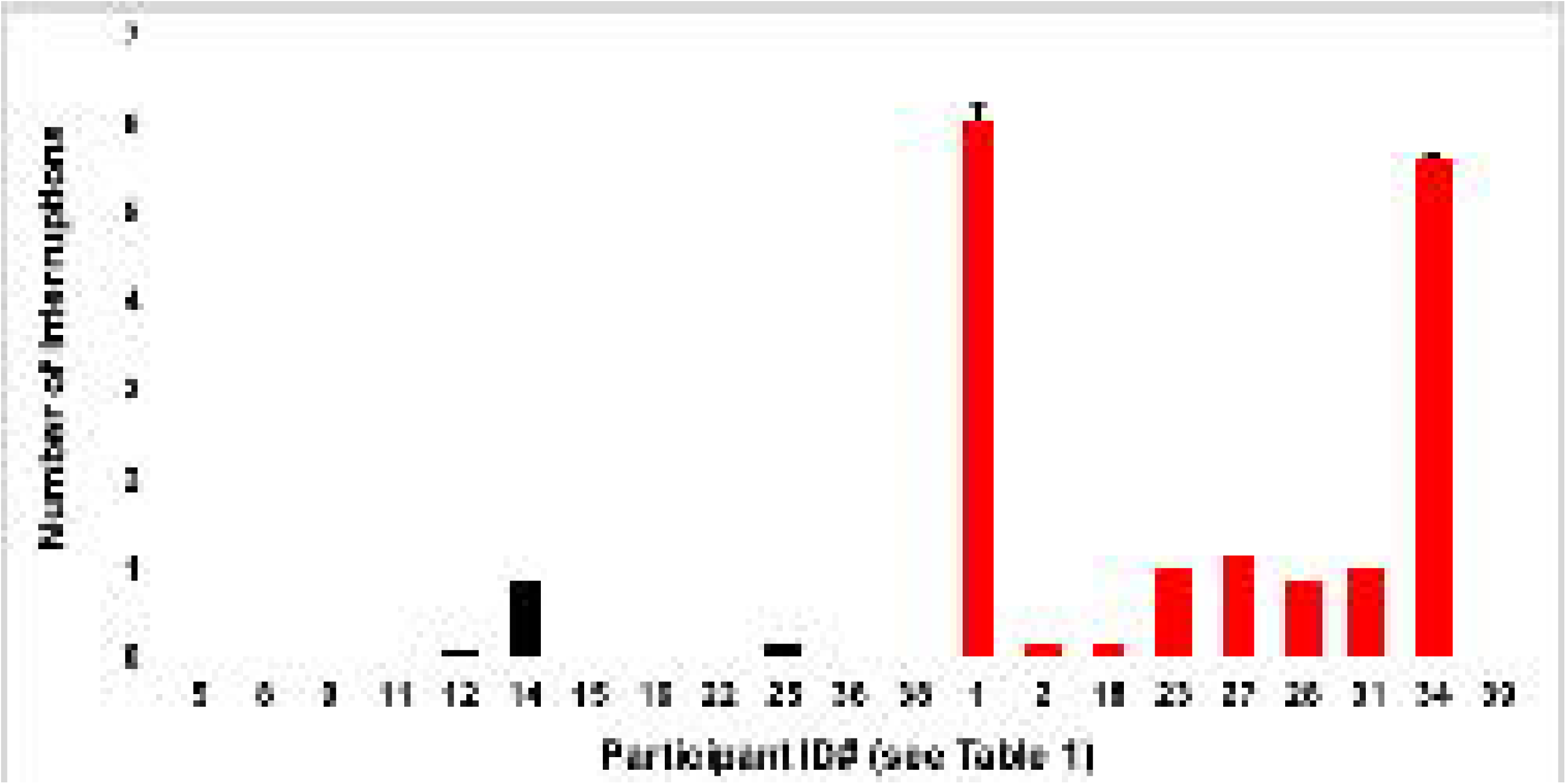
SP interruptions by group and participant. The average number of interruptions per session for each participant. Error bars represent S.D.

## 4 Discussion

The current study measured corticospinal excitatory and inhibitory pathways as reflected by MEP amplitude and SP duration, respectively, in individuals with and without SCI. Our data confirm the well-known finding that SCI individuals have higher TMS motor thresholds and lower MEP amplitudes than able-bodied volunteers, indicating reduced excitability of the motor cortex, reduced corticospinal transmission, reduced excitability of local spinal circuitry, or any combination thereof. Due to large variability *between* individuals, we found no significant difference in SP duration between SCI and able-bodied participants. Importantly, variability *within* individuals of SP duration is lower than variability of MEP amplitude, suggesting that SP duration may be a useful outcome measure with higher signal-to-noise ratio.

Appropriate movement requires inhibitory neural feedback as well as neural excitation. Consequently, imbalance between neural excitation and inhibition may contribute to the failure in restoration of useful motor function after SCI. Whereas MEP amplitude is usually regarded as an excitatory readout and SP duration as an inhibitory readout, both measures reflect a complex relationship between excitatory and inhibitory neurophysiology and clinical pathophysiology. A prolonged SP has been reported in a number of cerebral pathologies such as stroke^9^, epilepsy^10^, and depression^11^. On the other hand, shortened SP duration has been reported in bipolar disorder^12^, Parkinson’s^21^, Alzheimer’s disease^22^, and chronic neuropathic pain^13^.

Thus, lesions affecting the cortex, corticospinal tract, or segmental spinal circuits may affect SP duration in opposing fashion. Further complicating the interpretation of SP duration, a variety of experimental factors affect SP. In our study, we examined SP duration in the context of a maximal-effort pincer grip between the thumb and third finger, with a relatively low TMS intensity (110% of RMT). SP duration correlates positively with increases in TMS intensity, but does not correlate with level of volitional contraction^3,23^. SP duration is also highly task dependent. In young adults, the fractionated task of abducting the index finger demonstrated longer SP duration than a task involving pincer grip between the thumb and index finger, which demonstrated longer SP duration than a task involving power grip^24^. This suggests an inverse correlation between SP duration and the number of muscles used for a task^4^. Another study in healthy adults showed that increasing amounts of sensory afferent input shortened SP duration^25^. These studies demonstrated the importance of afferent feedback in downwardly modulating SP duration, presumably by inhibiting the cortical GABAergic interneurons that mediate SP^15^. Thus, SCI would be expected to prolong SP duration due to reduced afferent feedback through the lesioned cord.

Our study did not show a significant difference in SP duration between SCI and AB participants. We speculate that the relatively low TMS intensity used in our study contributed to shorter SP duration and increased variability between individuals, reducing sensitivity to detect differences^20^. Prior investigation of SP in the SCI population has been limited. In six participants after thoracolumbar SCI, SP duration in hand muscles above the lesion increased (APB muscle, “gentle” pincer effort; TMS intensity at 140-180% RMT), MEP amplitudes decreased, and cortical motor map representations shifted^14^. The altered cortical maps in this study, as well as the observed changes in muscles rostral to the spinal lesion level, suggested that these neurophysiological changes after SCI were cortical in origin.

Likewise, a study of 16 participants with chronic cervical SCI relative to 18 uninjured controls showed prolonged SP duration (FDI muscle, 25% effort, TMS intensity set to produce similar MEP amplitude across participants) after SCI, regardless of oral baclofen intake^7^. The difference in SP duration seen by Barry and colleagues was observed when evoked by TMS but not when evoked by transcranial electrical stimulation (TES) – since TMS activates corticospinal neurons indirectly through cortical interneurons, this discrepancy between TMS and TES pointed to cortical mechanisms for SP elongation after SCI. A study of nine males with chronic cervical SCI observed prolonged SP (extensor digitalis communis muscle, 10% effort, TMS intensity at 110% AMT), increased motor thresholds, and shifted cortical maps relative to uninjured controls^16^. Participants with greater spinal cord atrophy in that study showed relatively greater changes in SP and motor thresholds, suggesting that corticospinal transmission and local cord circuitry contributed to the observed neurophysiological changes.

On the contrary, a study of three individuals with cervical SCI showed loss of SP in two of three hand muscles and all three foot muscles (50% effort, TMS intensity at 100% of stimulator output)^17^. The authors speculated that SP loss derived from abnormal ascending sensory activity leading to cortical hyperexcitability, and/or increased local spinal neuronal excitability.

Our findings of decreased coefficients of variation in SP duration relative to MEP amplitude support the contention that SP duration may be a more reliable TMS metric than MEP amplitude. One other study measured MEPs and SP across time in individuals with chronic incomplete tetraplegia^18^. In that study, SP duration had relatively high reliability for both stronger and weaker muscles, whereas MEP amplitude had medium reliability for stronger muscles and poor reliability for weaker muscles. Studies in able-bodied participants provide further evidence that SP duration is more reliable across sessions than between individuals^20^. Importantly, resting motor threshold, which is determined at the beginning of any experimental TMS session, showed the least between-session variability of the measures we examined. Resting motor threshold demonstrated the lowest variability of multiple TMS measures in a study of elderly individuals with and without stroke^26^. It remains to be determined whether motor threshold is more sensitive than other measures to detecting changes after interventions aimed to increase central neural transmission in people with SCI.

We observed an overwhelmingly higher prevalence of SP ‘interruption’ frequency in SCI relative to AB participants. These short (<20 ms) periods of EMG activity in the midst of the SP appear similar to ‘late excitatory potentials (LEP)’ noted by Wilson et al^27^ and ‘breakthrough EMG activity’ discussed by Hupfeld at al^4^. Intra-SP EMG activity has been hypothesized to originate from two sources: muscle spindle-gamma motoneuron reflex activity in response to muscle lengthening during the SP; and transient cortical disinhibition^27^. Caudal to the lesion, increased spindle-mediated reflex activity is well documented after SCI. At the lesion, fasciculations of individual motor units represent spontaneous hyperexcitable discharges in upper extremity muscles commonly seen after cervical SCI^28^. Therefore, it is not surprising that SCI participants demonstrated a much higher SP interruption rate than AB participants.

Our study has several important limitations: Due to time constraints, only 5-6 SP trials were collected per session, less than the roughly 20 trials needed to maximize reliability of most TMS measures^4,29^. The SCI group was older than the control group. A majority of past studies suggest that age negatively correlates with SP duration, which would mean that the results of this study may underestimate the true difference in SP duration between SCI and non-SCI groups^4^. Individuals in the SCI group were more likely to be taking oral baclofen or other neural inhibitory medications. However, unlike intrathecal baclofen, oral baclofen has not clearly been shown to affect SP duration^7,8^. Furthermore, individuals with SCI have obviously lower ability to contract target muscles than non-SCI participants during SP elicitation. However, this would be the case whether at maximal effort or at any set percentage of an individual’s volitional effort, and muscle contraction intensity does not play a major role in SP duration regardless^3,23^. TMS intensity during SP measurements was at 110% of resting motor threshold – most studies have used higher TMS intensities, which is a key factor in longer SP duration and may reduce intersession variability^3,20,23^.

## Conclusion

In individuals with chronic cervical SCI relative to able-bodied controls, we confirmed the well-known findings that SCI individuals have lower TMS evoked potential amplitudes and a tendency toward higher TMS motor thresholds. We did not observe significantly longer SP duration in SCI individuals. Importantly, we observed significantly lower within-person variability of SP duration than within-person variability of TMS response amplitude, suggesting that SP duration may be a useful outcome measure with higher signal-to-noise ratio. Ongoing and future studies in our lab will further investigate silent periods induced by either cortical or cutaneous stimulation^30^ to correlate EMG with electroencephalographic features and shed more mechanistic insight into these phenomena.

## Supporting information

Supplemental data by Subject

Supplemental Data by Session

## Data Availability

All relevant data are within the manuscript and its Supporting Information files.

## Data Archiving

Individual-level data is included as Supplementary Data 1 (by participant, averaged across sessions) and Supplementary Data 2 (by participant per session). All deidentified data is freely available to any investigator upon request.

## Statement of Ethics

All procedures were approved by the Institutional Review Board of the James J. Peters VA Medical Center, Bronx, NY (HAR-15-001). We certify that all applicable institutional and governmental regulations concerning the ethical participation of human volunteers were followed during the course of this research.

## Author Contributions

HJS: Data acquisition; data interpretation; manuscript drafting and revision

JRW: Data acquisition; data interpretation; manuscript drafting and revision

OA: Data interpretation; manuscript revision

YKW: Data acquisition; data interpretation; manuscript revision

NYH: Study conception and design; data interpretation; manuscript drafting and revision.

## Legends

**Supplementary Data 1 – Participant-level data averaged across sessions**. InterssNum, number of interruptions per session; CVSPdurIntra, intrasession coefficient of variation for SP_DUR_; CVMEPrestIntra, intrasession coefficient of variation for MEP_REST_; CVMEPactiveIntra, intrasession coefficient of variation for MEP_ACTIVE_; CVRMTInter, intersession coefficient of variation for RMT; CVSPdurInter, intersession coefficient of variation for SP_DUR_; CVMEPrestInter, intersession coefficient of variation for MEP_REST_; CVMEPactiveInter, intersession coefficient of variation for MEP_ACTIVE_.

**Supplementary Data 2 – Participant-level data listed per session**.

